# A qualitative process evaluation using the behaviour change wheel approach: Did a whole genome sequence report form (SRF) used to reduce nosocomial SARS-CoV-2 within UK hospitals operate as anticipated?

**DOI:** 10.1101/2022.08.30.22279427

**Authors:** Paul Flowers, Ruth Leiser, Fiona Mapp, Julie McLeod, Oliver Stirrup, Christopher JR Illingworth, James Blackstone, Judith Breuer

## Abstract

To conduct a process evaluation of a whole genome sequence report form (SRF) used to reduce nosocomial SARS-CoV-2 through changing infection prevention and control (IPC) behaviours. Here using qualitative behavioural analyses we report how the SRF worked.

**Methods:** Prior to a multisite non-randomised trial of its effectiveness, the SRF was coded in relation to its putative behaviour change content (using the theoretical domains framework (TDF), the behaviour change wheel (BCW) and the behaviour change technique taxonomy (BCTTv1)). After the SRF had been used, through the peak of the Alpha variant, we conducted in-depth interviews from diverse professional staff (N=39) from a heterogeneous purposive sub-sample of hospital trial sites (n=5/14). Deductive thematic analysis explored participants’ accounts of using the SRF according to its putative content in addition to inductive exploration of their experiences.

**Results:** We found empirical support for the putative theoretical mechanisms of ‘Knowledge’ and ‘Behavioural regulation’, as well as for intervention functions of ‘Education’ and ‘Persuasion’ and ‘Enablement’, and for particular BCTs ‘1.2 Problem solving’, ‘2.6 Biofeedback’, ‘2.7 Feedback on outcomes of behaviour’, and ‘7.1 Prompts and cues’. Most participants found the SRF useful and believed it could shape IPC behaviour.

**Conclusions:** Our process evaluation of the SRF provided granular and general support for the SRF working to change IPC behaviours. Our analysis highlighted useful SRF content. However, we also note that, without complementary work on systematically embedding the SRF within routine practice and wider hospital systems, it may not reach its full potential to reduce nosocomial infection.

**What is already known on this subject?:**

- Health psychology remains under-exploited within infection prevention and control (IPC) interventions
- For genomic insights to be understood by a range of health care professionals and elicit changes in IPC behaviour, ways of translating complex genomic insights into a simple format are needed. These simple translation tools can be described as whole genome sequence report forms (SRFs)
- Nothing is currently known about the use of SRFs, for SARS-CoV-2 or other infections, to change hospital-based IPC behaviour.
- Health psychological tools such as the behaviour change wheel (BCW), the theoretical domains framework (TDF), and the behaviour change technique taxonomy (BCTTv1) are widely used to <u>develop</u> behaviour change interventions but are rarely used to <u>evaluate</u> them
- Contemporary guidance on conducting process evaluations highlights the value of explicitly theorising <u>how</u> an intervention is intended to work before systematically examining how it actually worked in practice

**What does this study add?:**

- The paper presents a novel worked example of using tools from health psychology within a qualitative process evaluation of using an SRF during the COVID-19 pandemic in UK hospitals
- This paper is the first to report how people experienced using whole genome sequence report forms (SRFs) in order to change hospital-based IPC behaviour
- We provide qualitative evidence detailing empirical support for much of the SRF’s putative content, including casual mechanisms ‘Knowledge’ and ‘Behavioural regulation’, intervention functions such as ‘Education’ and ‘Enablement’, and for particular BCTs: ‘1.2 Problem solving’, ‘2.6 Biofeedback’, ‘2.7 Feedback on outcomes of behaviour’, and ‘7.1 Prompts and cues’

## Introduction

Nosocomial infection of SARS-CoV-2 where transmission occurred within hospitals - was a major problem throughout the COVID-19 pandemic, as it presented significant health risk to both patients and healthcare workers (Abbas et al., 2021; Lucey et al., 2021; Oliver, 2021 and Read et al., 2021). Nosocomial COVID-19 added to the longstanding problem of healthcare-associated infections (HCAIs) (Haque et al, 2018).

Whole genome sequencing (WGS) can be insightful for changing infection prevention and control (IPC) behaviour, for example, by providing incremental insights into infectious disease transmission within healthcare (Harris et al., 2013; Quick et al., 2016; Van El et al., 2013). In an asynchronous way, WGS per se can inform IPC practice by providing insights into historical transmission routes and their relation to past IPC behaviour (e.g., enhanced cleaning, patient isolation, patient movement or visitor restrictions, personal protective equipment, contract tracing). Prior to COVID-19, there was growing debate about the potential of WGS for assisting with reducing HCAIs (Balloux et al., 2018; Peacock et al., 2018). However, WGS had not been used to synchronously (in real-, or near real-time) to change IPC behaviour and many factors inhibited this application (Balloux et al., 2018; Parcell et al., 2021). In particular: (1) the substantial infrastructure required, (2) the political, professional, and personal will to trial it at scale, and (3) the complexity of the insights typically delivered through WGS, which require technical and expert understanding, negatively impacting its cost-effectiveness. However, the scale of the UK’s response to the COVID-19 pandemic removed many of these long-standing barriers simultaneously. WGS of SARS-CoV-2 became a vital global surveillance tool (e.g., identifying variants of concern), rapid investment across UK hospitals and laboratories provided the necessary WGS infrastructure (Blackstone et al., 2022). Equally - unlike any other preceding time period - governments, healthcare professionals (HCPs), and researchers were all galvanised to act on WGS insights because of the COVID pandemic and high levels of nosocomial infection. However, the necessity for expertise to interpret and understand WGS output remained problematic. In this context, the COG-UK Hospital Onset COVID-19 Infection (HOCI) study (Blackstone et al., 2022) offered an opportunity to examine the effectiveness of rapid (<48 hour) WGS reporting to shape IPC behaviour and reduce nosocomial COVID-19 infection (Stirrup et al., 2022). For the HOCI study, a bespoke sequencing report form (SRF) was designed to translate WGS insights into comprehensible and actionable insights for IPC teams. If viral samples could be collected and processed rapidly and efficiently, the SRF then provided near real-time identification of likely cases of nosocomial transmission, facilitating HCPs to take appropriate IPC action.

To date, health psychological approaches remain under-used within interventions to reduce HCAI, although their potential benefits have been widely articulated (e.g., Edwards et al., 2012; Greene et al., 2022; Price et al., 2018; Price et al., 2018; von Lengerke et al., 2019). While the HOCI study was not unique in examining the real-time use of sequence data to address nosocomial transmission (Illingworth, 2021), parallel studies have not been accompanied by comprehensive evaluation of the reporting tools designed to change behaviour. Given its centrality within the HOCI study, and its potential to be used to manage future infections beyond SARS-CoV-2 we believed the SRF used within the HOCI study merited systematic investigation.

### Aims

To conduct a process evaluation of the SRF to explore if it worked as anticipated and how people reported using it in practice.

### Research questions

1. What were the putative active ingredients of the SRF?
2. Was there evidence to support the SRF working as anticipated?
3. How did the SRF work in practice?

## Methods

### Design

A sequential, pre- and post-focussed qualitative process evaluation. Pre-trial work used documentary analysis and interviews with experts. Post-trial work used semi-structured qualitative interviews with diverse HCPs and inductive & deductive thematic analysis (Braun & Clark, 2006). These two stages of data collection and analysis are detailed below.

Guidance exists for conducting process evaluations (e.g., Moore et al., 2015). It is intended to help with identifying what has worked and what has not within interventions. Such guidance typically stresses the centrality of theorising intervention content, preferably prior to intervention roll-out as well how interventions work in practice, as well as understanding intervention context (Craig et al., 2018). Although rare in health psychology process evaluations are more common within implementation science (e.g., Curran et al., 2013; Kislov et al., 2019; May et al., 2018) and there is increasing recognition of the advantages of deeper knowledge transfer between these sister disciplines (Presseau et al., 2022).

#### 1. Pre-trial (initial analysis of the how the SRF should work)

Details of the full approach to process evaluation are available elsewhere (Flowers et al., 2021). Here, in brief, we provide an overview of the pre-trial work pertinent to the focused analysis of the SRF reported here.

##### Pre-trial data

Independent of the wider interdisciplinary team directly involved in the development of the SRF, a Health Psychologist (PF) analysed a purposive heterogeneous sample of completed SRFs (see Figure 1 for example). This was complemented by informal interviews (N=9) with a range of experts with an interest in nosocomial infections (e.g., IPC staff, virologists, microbiologists).

**Figure 1.**
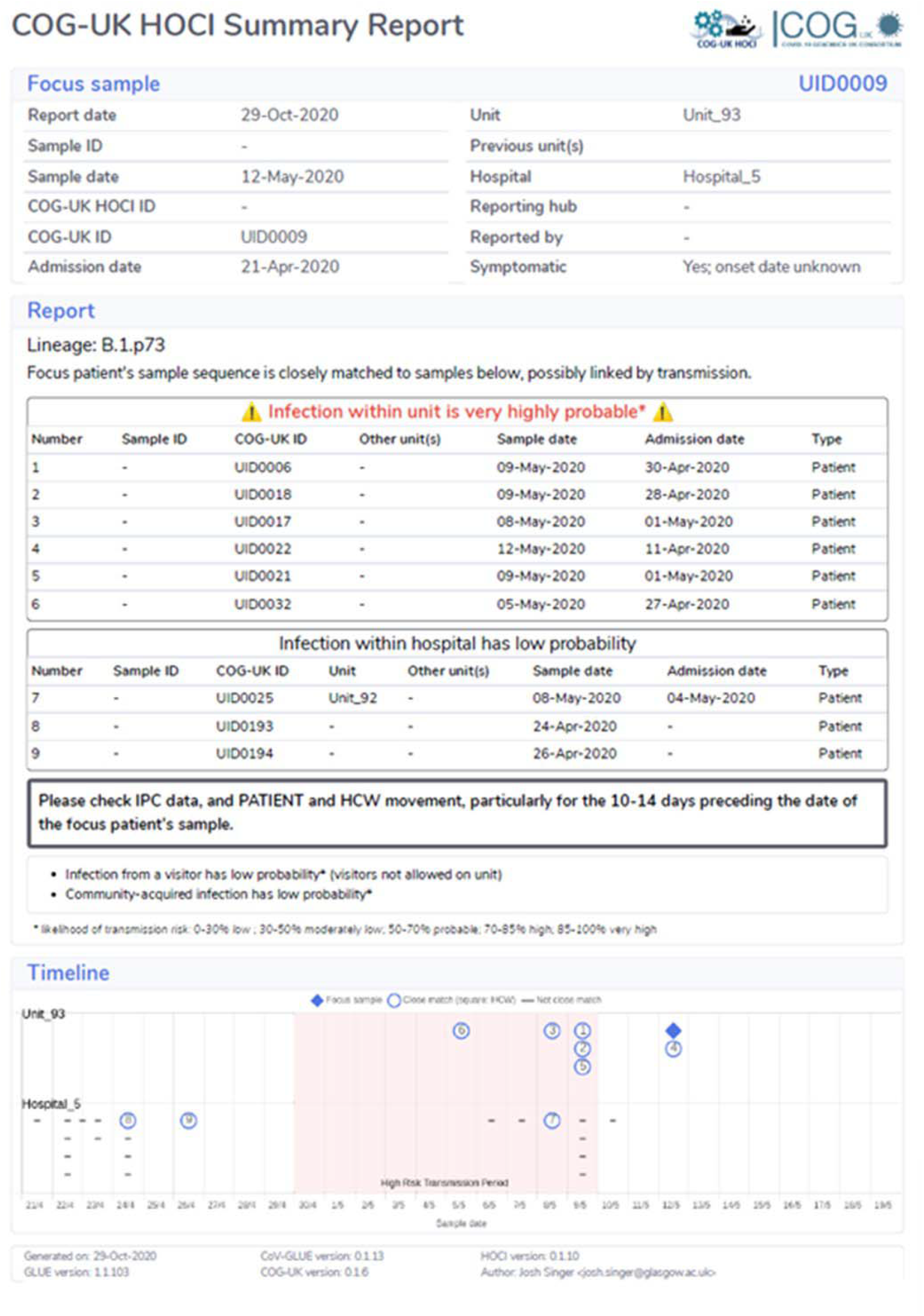
An example of the sequence report form.

##### Pre-trial analysis

Analysis for the first research question incorporated a series of tools which, in combination, described the putative active ingredients of the SRF and their theoretical underpinnings. PF analysed the SRF using the theoretical domains framework (TDF) (Cane et al., 2012) to detail putative theoretical mechanisms; the behaviour change wheel (BCW) (Michie et al., 2011) to detail its ‘intervention functions’; and the behaviour change technique taxonomy (BCTT, Michie et al., 2013) to understand its behaviour change content. Analysis was audited by FM, RL and JM, and shared with the wider team. Analysis of the SRF’s content (RQ1) categorised elements as either having a major or minor role. Major mechanisms were those that the team considered most explicit and clearly obvious to the function of the SRF. Minor mechanisms were those which were more implicit, less obvious, and had a secondary role within the SRF.

#### 2. Post-trial (qualitative analysis of how the SRF did work)

##### Sampling

From 14 total study sites, a purposive sample of five focal sites were selected for in-depth data collection. To show heterogeneity of experience with the SRF, sites were selected to be varied in relation to prior case rates, hospital size, familiarity with sequencing, and geography. Data collection started after the SRF had been used for at least 14 days within the rapid phase of WGS – where the target turnaround time for output was within 48hrs.

##### Recruitment

Within each site, a senior member of staff involved in the study approached a broad range of professionals involved in the use and implementation of WGS. Those interested in participating were sent participant information sheets. A mutually convenient time was arranged, and interviews were conducted using online meeting platforms. One researcher (FM) conducted all interviews.

##### Participants

Within each of the selected sites, a sample of between six and nine participants took part. The final sample comprised 39 participants (n=9 site 1; n = 7 site 2; n=8 site 3; n=8 site 4; n=6 site 5), 27 identified as female (69%) and 12 as male (31%), with an age range of 20-70. Participants’ roles within the study varied and were not limited to those who directly used the SRF to change IPC behaviour (e.g., clinical fellow, sequencing lab manager, bioinformatician, and research nurse).

##### Post-trial data collection

Sites were enrolled in this study between October 2020 and April 2021. Data collection occurred between December 23rd 2020 and June 2nd 2021, across the peak and decline of the Alpha variant. A topic guide was used, exploring participants’ thoughts and experiences of the SRF in one-to-one interviews (30-90 minutes). Data collection included a focused discussion on the SRF, including collecting information on what people did or did not find useful about it. Interviews were audio recorded, transcribed by a professional transcribing company, and anonymised.

##### Post-trial data analysis

Analysis was an iterative process involving cycles of both deductive and inductive thematic analysis. First, PF and FM engaged in multiple data readings and discussions. After this, an initial coding frame was developed. This contained broad categories of data, some of which were pre-specified (i.e., participant perspectives on the SRF), others were identified from the data. A wider team of five researchers including PF, JM and FM then became responsible for one site each, and used the coding-frame. The data from each of the sites were then collated. Further inductive, and then deductive, analyses were conducted (led by PF). These latter analyses focused on deductively mapping participant data in relation to our ideas about the putative content of the SRF (RQ2) - this meant systematically mapping participant data to the TDF domains, intervention functions, and individual BCTs - and inductive analysis of the participants’ perspectives on the SRF (RQ3). All analyses were audited closely by one other analyst (RL) trained in the use of the TDF, BCW, and BCTT, in addition to iterative discussions with the wider interdisciplinary team.

Post-trial analysis of the SRF categorised the relative level of support for the putative behaviour change content of the SRF within the qualitative data as either ‘strong’, ‘weak’, ‘nuanced’ or ‘no support’. Relative support was gauged primarily by frequency of data occurrence, both across the interviews as a whole, and within each participant’s account. Beyond frequency, relative support was assessed by the pragmatic importance of the finding, the temporal and historical context of the data (e.g., in relation to the peak of the Alpha variant). Iterative discussion within the research team finalised the agreed level of support.

### Ethical approval

Ethical approval was given by Cambridge South Research Ethics Committee (20/EE/0118).

## Results

### RQ1: Pre-trial – what were the putative active ingredients of the SRF?

In this section we describe how, before the trial and interview data collection began, we anticipated how the SRF would work. The second column in Tables 1, 2 and 3 all detail our pre-trial analysis of the SRF content.

**Table 1.**
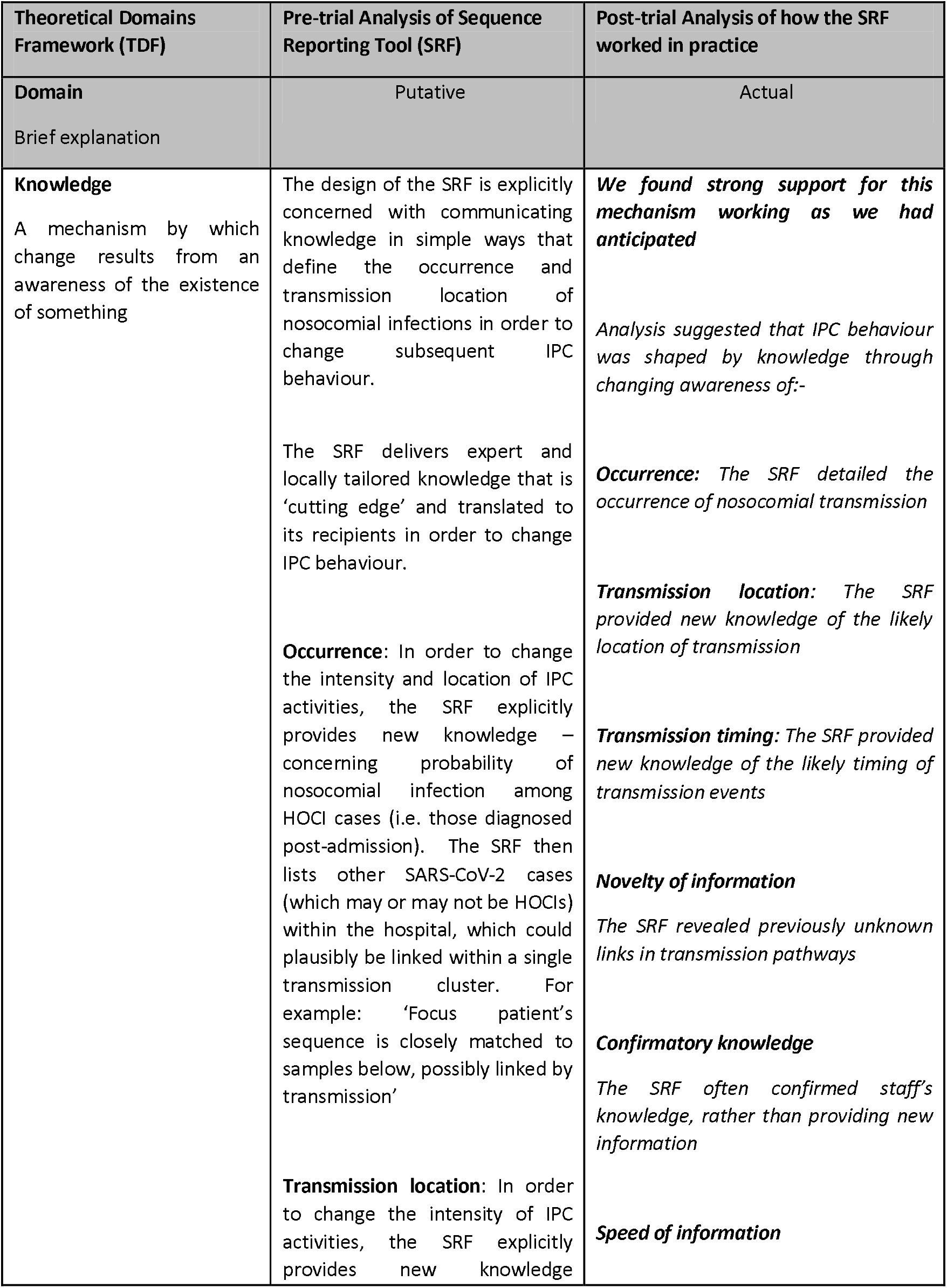
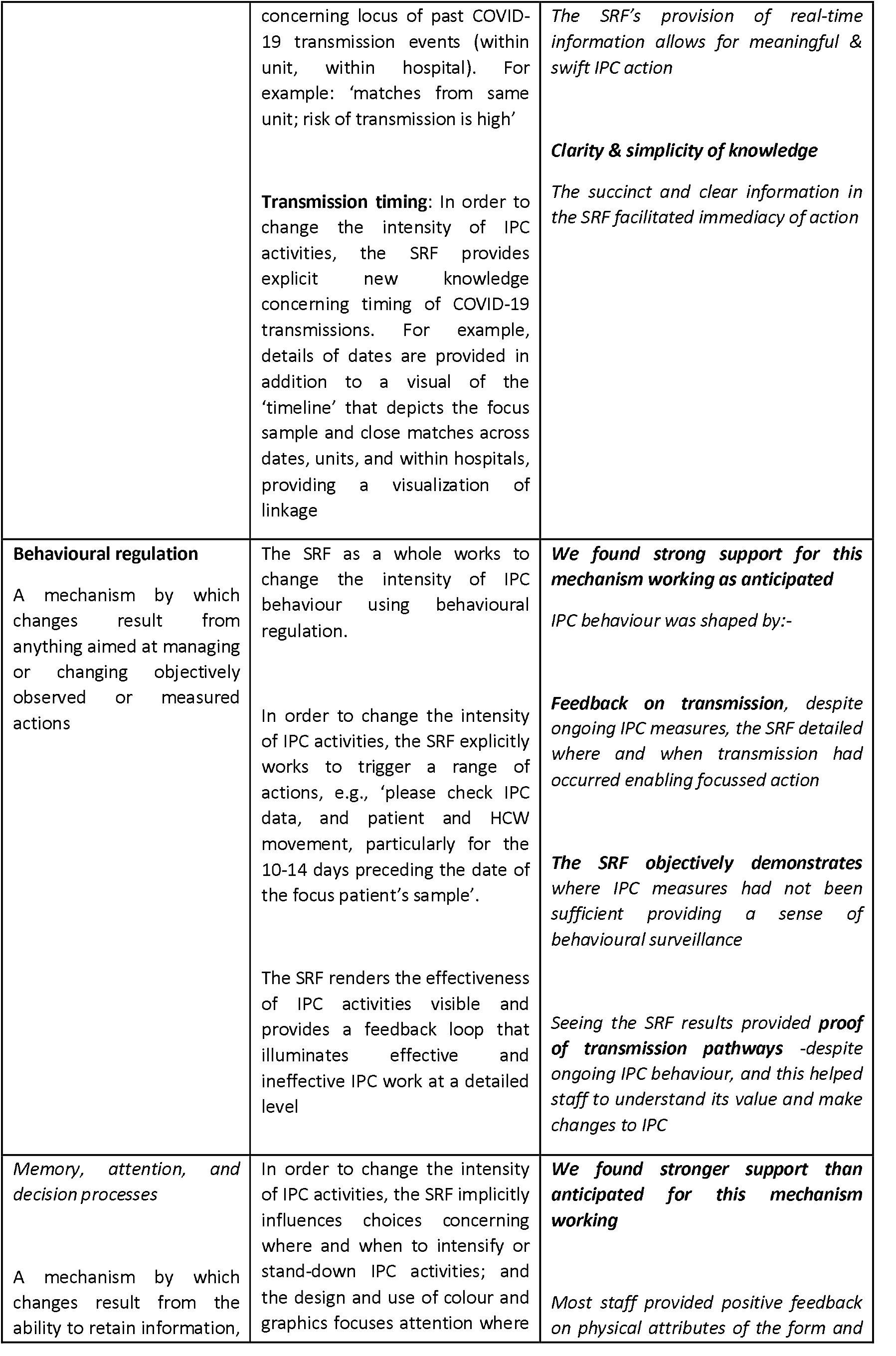
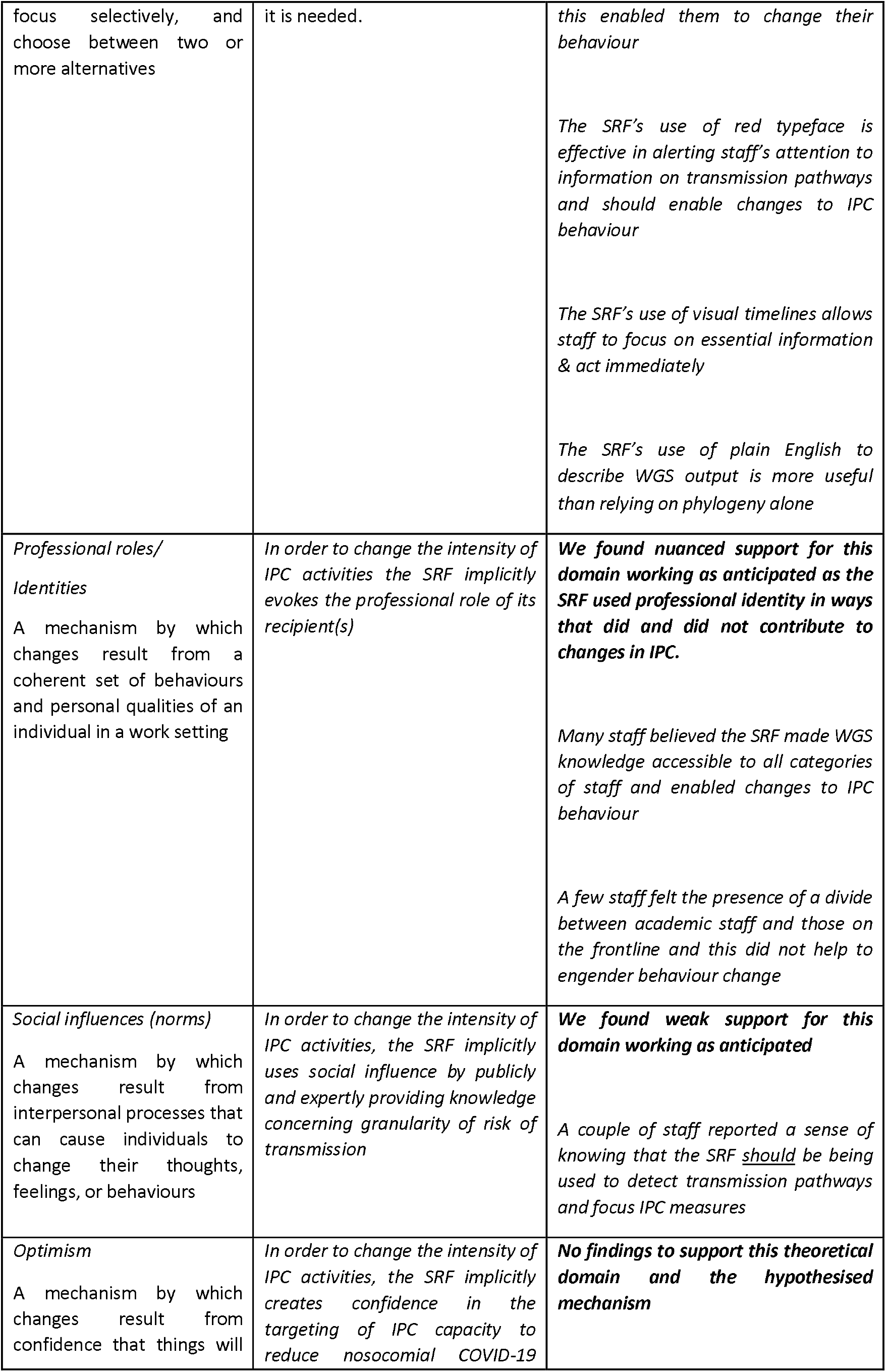
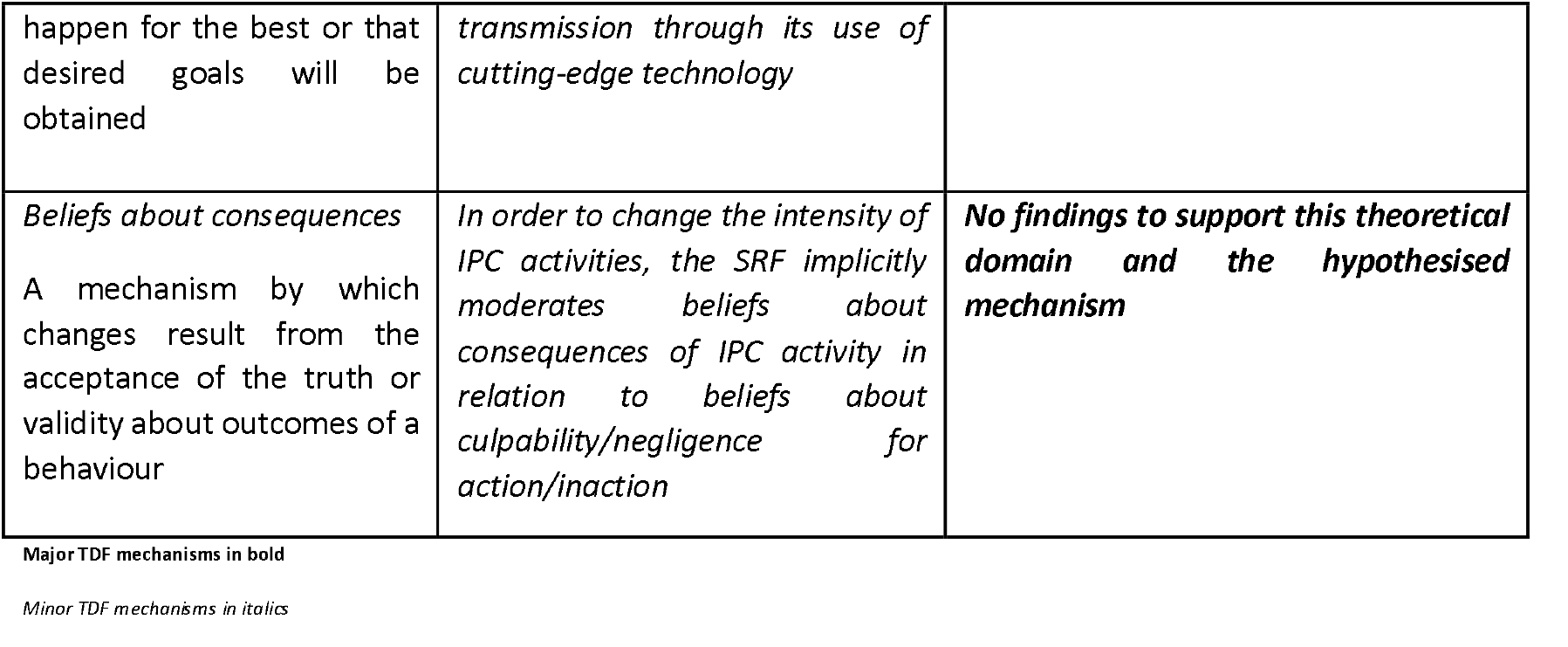
Putative theoretical mechanisms and their empirical support. Domains in bold refer to those identified as being major mechanisms

**Table 2.** Putative intervention functions and their empirical support.

| **Intervention functions & definition** | **Pre-trial analysis of Sequence Reporting Tool (SRF)** | **Post-trial analysis of how the SRF worked in practice** |
| --- | --- | --- |
| **Education**<br>Increasing knowledge or understanding | In order to change the intensity of IPC activities, the SRF explicitly provides new knowledge and understanding of HOCIs | ***We found strong support for this intervention function***<br>*The SRF provides the knowledge to take swift & effective action, by allowing people to understand transmission pathways better and make changes to IPC*<br>*The SRF sometimes confirmed what staff already knew, or simply reassured them that their efforts were worthwhile but did sometimes lead to standing down of ongoing IPC behaviours* |
| **Enablement**<br>Increasing means/reducing barriers to increase capability | In order to change the intensity of IPC activities, the SRF clearly minimizes uncertainty to reduce barriers to stimulate focused IPC action | ***We found strong support for this intervention function***<br>*The SRF provided a sense of clarity and the capacity* |
| or opportunity |  | *for understanding output at a glance, allowing for immediacy of IPC response*<br>*The SRF allowed staff to carry out duties more efficiently and the SRF's simple, effective design facilitated its use* |
| **Persuasion**<br>Using communication to induce positive or negative feelings or stimulate action | In order to change the intensity of IPC activities, the SRF clearly uses communication of expert and locally tailored knowledge to stimulate changes in the intensity of IPC activity | ***We found nuanced support for this intervention function***<br>*The way by which the SRF communicated new information was well-received by some but not all; positive feeling towards the content of the SRF encouraged some staff to act* |
| **Modelling**<br>Providing an example for people to aspire to or imitate | In order to change the intensity of IPC activities the SRF provides a specific prompt to check IPC data and patient and HCW movement | ***No findings to support this intervention function*** |

Major intervention functions mechanisms in bold
Minor intervention mechanisms in italics

**Table 3.**
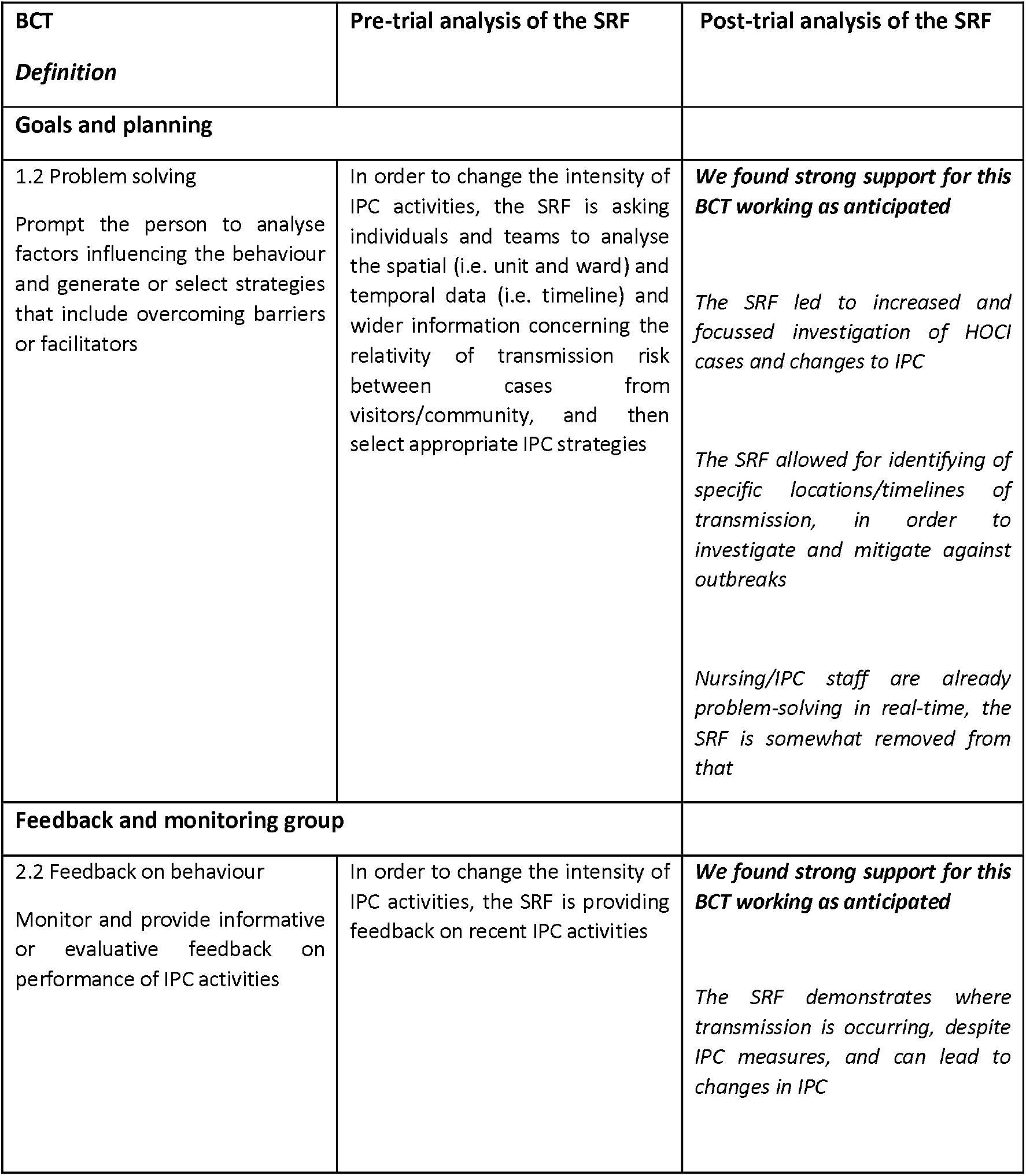
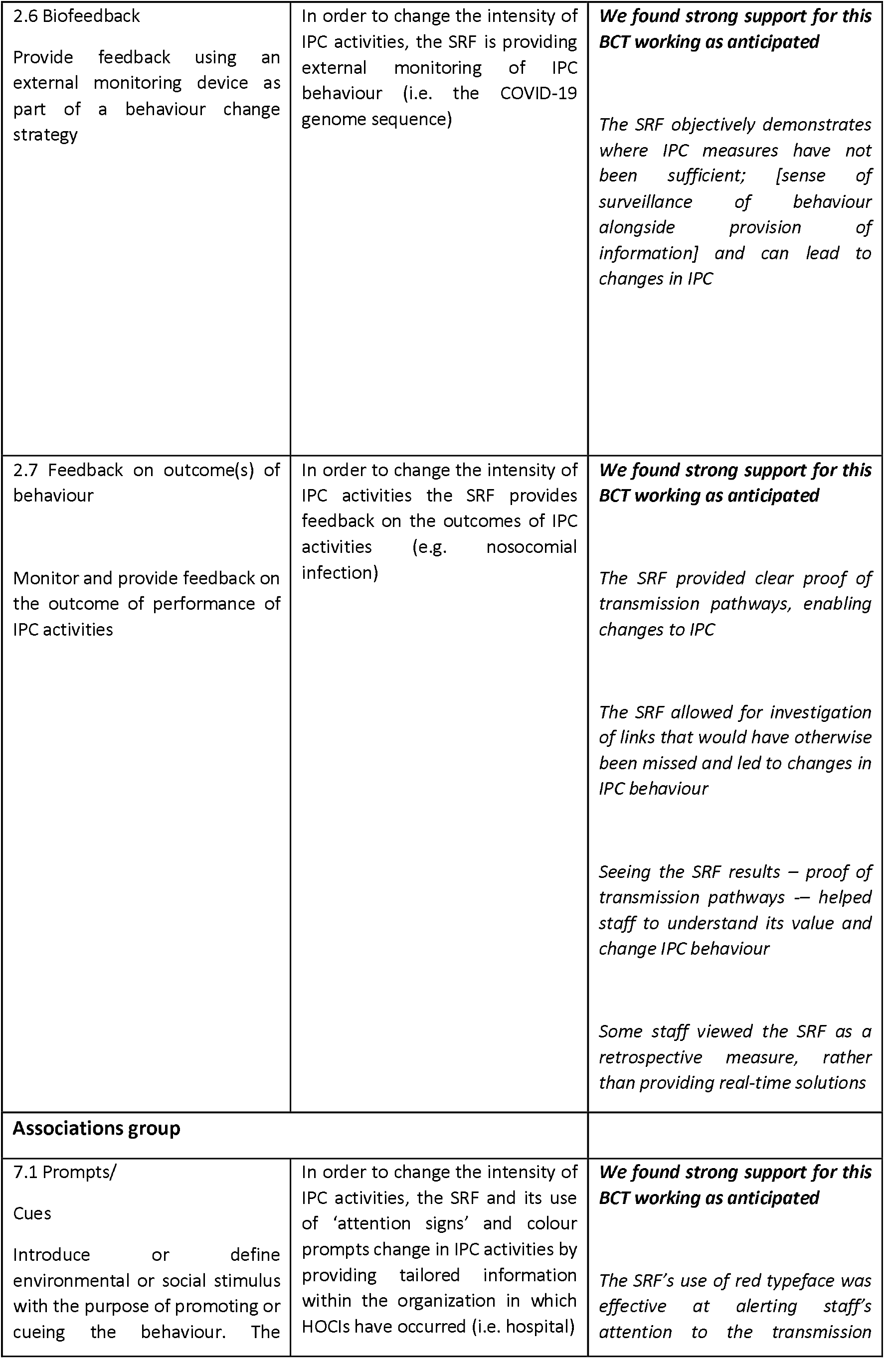
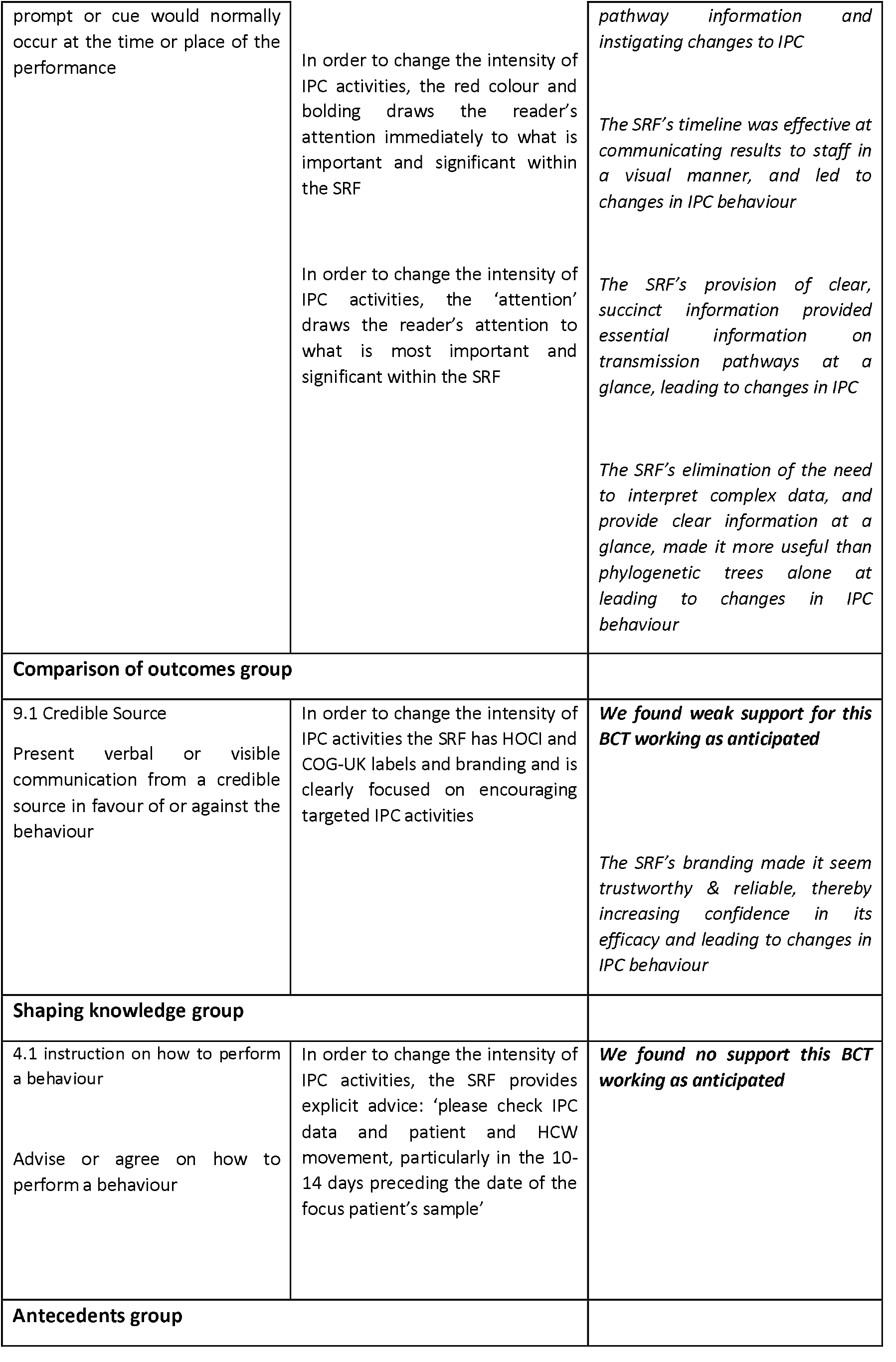
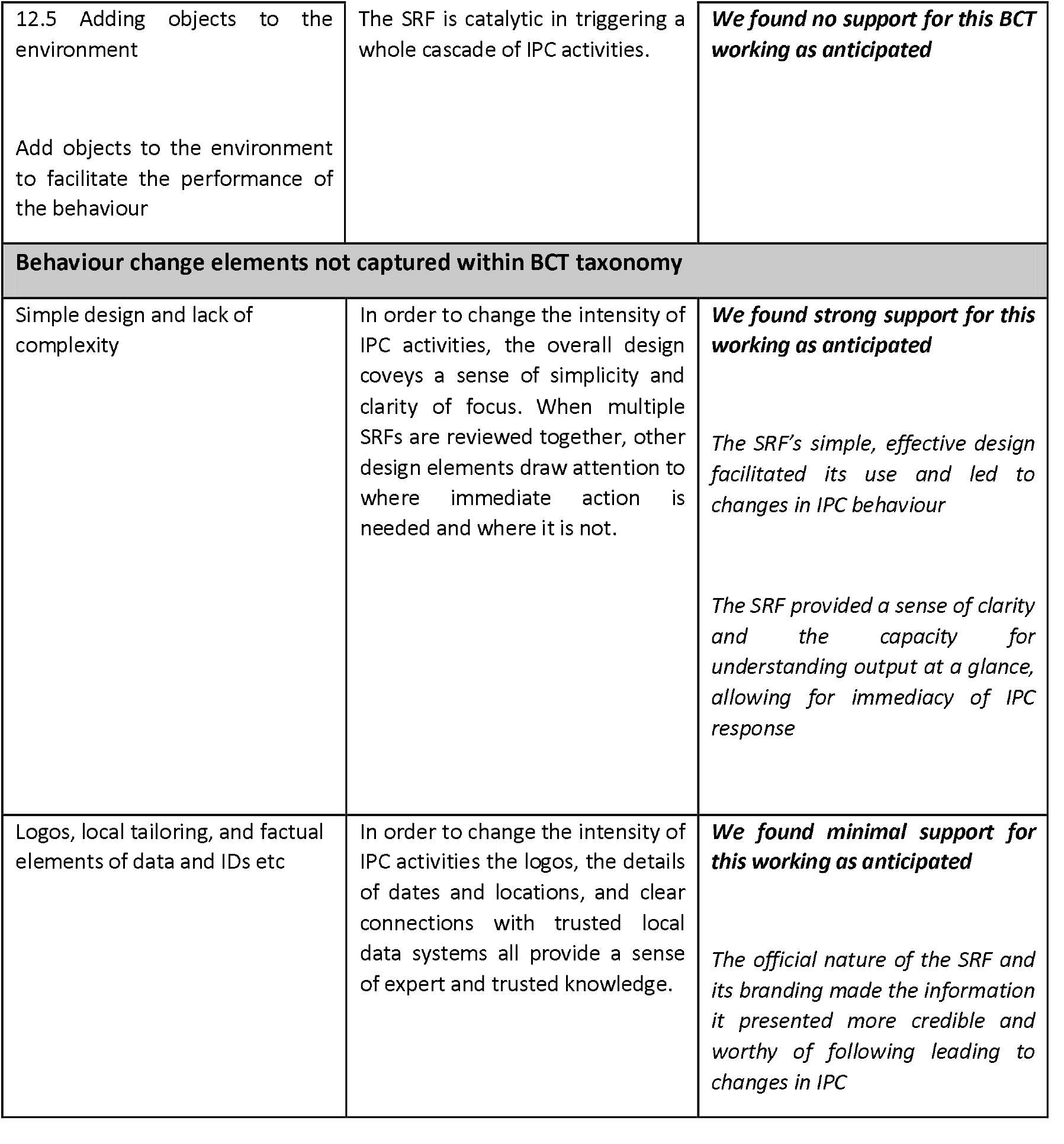
Relative support for the BCTS used within the SRF.

#### a) Theoretical mechanisms within the SRF

The TDF was used to capture the theoretical mechanisms underpinning the way we imagined the SRF might work to change the intensity and location of IPC behaviour. We identified two major and five minor domains (see Table 1). In relation to the two major domains, firstly, we highlighted that ‘Knowledge’ was a very important theoretical domain underpinning the SRF. The SRF was designed to provide rapid knowledge of nosocomial infection regarding its occurrence, transmission location, and transmission timing. Secondly, our analysis suggested that ‘Behavioural regulation’ was also a central major domain, as the SRF provides objective retrospective feedback concerning recent IPC behaviour, intended to change the intensity and location of future IPC behaviour.

A range of minor, causal mechanisms were also detailed as being present. These included ‘Memory, attention, and decision-making processes’ in which the SRF’s use of colour, graphics, and symbols gains recipient attention and elicits changes in IPC behaviour; ‘Professional role and identity’ whereby the SRF works by evoking the professional identities of its recipients and ideas of professional role and responsibility to change IPC behaviour; ‘Social influence’ whereby the public and expert knowledge provided by the SRF can be understood to be providing injunctive norms about the need for changes to IPC behaviour; ‘Optimism’ by which the use of cutting-edge technology (i.e. whole genome sequencing) reflected within the SRF encourages changes in the intensity and location of IPC behaviour; and finally, ‘Beliefs about consequences’ in which the IPC behaviours are changed by beliefs about culpability regarding IPC.

#### b) The SRF and its overarching intervention functions

Using the BCW, our analysis suggested that the SRF would use four intervention functions – two major intervention functions and two minor (see Table 2). In relation to the first major intervention function and relating to the SRF’s use of the theoretical mechanisms of ‘Knowledge’, we noted the SRF used ‘Education’ as a central way of changing the intensity and location of IPC behaviour. The SRF educated its recipients about the occurrence, transmission location, and the timing of transmission events. The second major intervention function identified in this pre-trial work was ‘Enablement’ in which uncertainty around nosocomial transmission is being reduced by the SRF, and this subsequently stimulates changes in the intensity and location of IPC behaviour.

Minor intervention functions identified included ‘Persuasion’ and ‘Modelling’. For ‘Persuasion’, the SRF works by communicating expert and locally tailored knowledge to change IPC behaviour. In relation to ‘Modelling’ the SRF can be seen to provide an example of what the recipient should aspire to in relation to the prompt to ‘check IPC data and patient and HCW movement’.

#### c) The SRF and its deployment of specific BCTs

Analysis suggested the SRF would use several BCT groups and individual BCTs to enact changes in the intensity and location of IPC activities (see Table 3). These are drawn from six of the sixteen behaviour change technique groups. Key groups included ‘Goals and planning’, ‘Feedback and monitoring’, ‘Associations’, ‘Comparison of outcomes’, ‘Shaping knowledge’ and ‘Antecedents’.

At the level of individual BCTs, our analysis highlighted that the SRF would work because it used the BCT ‘1.2 Problem Solving’. Therein, the provision of new knowledge concerning transmission, via the SRF’s ‘Education’ function, in addition to the explicit prompt within the SRF to ‘check data and patient movement’ all work to challenge the recipient to generate strategies for improved IPC. Equally, the cluster of related BCTs, ‘2.2 Feedback on behaviour’, ‘2.6 Biofeedback’, and ‘2.7 Feedback on outcomes of behaviour’ are all core parts of the SRF, as it is intended to change the intensity and location of IPC behaviour by providing external feedback on recent IPC behaviour and its relative performance. The SRF also uses the BCT ‘7.1 Prompts/Cues’ to change IPC behaviour by using attention signs and colour prompts. These draw the recipient’s attention to what is important and significant. Furthermore, within the SRF we believed that the BCT ‘9.1 Credible source’ was being deployed, as it used clear branding and encouragement to change IPC behaviour. We also coded the specific prompt of ‘please check IPC data and patient and HCW movement, particularly in the 10-14 days preceding the date of the focus patient’s sample’ as ‘4.1 Instructions on how to perform a behaviour’. Finally, we also coded the form itself as ‘12.5 Adding objects to the environment’, as we thought it plausible that the SRF could be considered a catalytic object, in and of itself, that could change the intensity and location of IPC behaviour.

### RQ2: Post-trial – was there evidence to support the SRF working as anticipated?

In this section we describe how, on the basis of our post-trial analysis of the interview data, we determined if the SRF had actually worked in practice as expected. The third column in Tables 1, 2 and 3 all summarise this post-trial analysis of the SRF content.

#### a) Understand how the SRF worked through considering its theoretical mechanisms

Table 1 shows that, in relation to theoretical mechanisms, our analysis of participant data provided broad support for the SRF working as anticipated. With regard to the two main mechanisms (e.g., ‘Knowledge’ and ‘Behavioural regulation’) that we had previously identified as particularly important within the SRF, we found strong support.

We found the SRF’s provision of knowledge relating to occurrence, transmission location, and timing did drive changes in the intensity of IPC behaviour. However, when this knowledge was delivered rapidly, it was seen as novel, confirmed clinical hunches, or supported actions already made, it was understood to be particularly important: ‘…they [SRFs] were gems, gems of information that could just be sent off’ (Site 5). For behavioural feedback, there was also strong support for this mechanism working as anticipated: ‘we’ve been able to use it to feed back to staff’ (Site 1)

In relation to the mechanisms that we had previously identified as having a lesser role in the way the SRF worked, we found stronger support for ‘Memory, attention, and decision-making processes’ than we had anticipated: ‘this infection within unit is very highly probable’, it’s a really clear, straightforward message. I like the red and the sort of warning signs, because I know that people sort of skip things’ (Site 1). We found quite nuanced support for ‘Professional roles/identity’; although the report spoke to a range of professional identities, for some staff working solely on the front line, it was not seen as speaking to their job role and the implications that entailed. Equally, contrary to our initial coding, we found minimal support for ‘Social influences’. Finally, for ‘Optimism’ and ‘Beliefs about consequences’ we found no support at all.

#### b) Understand how the SRF worked through considering its broad intervention functions

In relation to analysis supporting the SRF’s use of the BCW’s putative intervention functions, Table 2 describes how we found strong support for both ‘Education’ and ‘Enablement’. We also found some nuanced support for ‘Persuasion’, yet no support for ‘Modelling’.

Findings supported pre-trial analysis which identified Education and Enablement as central intervention functions. The importance of Education was anticipated partly due to the predicted importance of the TDF domain of Knowledge. It was clear how crucial increased understanding among staff would be to the SRF’s success, and this was reflected in the data. Our pre-trial analysis also established the theorised major role of Enablement. This intervention function was intended to actively circumvent barriers, thereby enabling staff to deploy IPC measures in the most effective and efficient way possible. The data showed support for this intervention function, particularly with regard to the SRF’s simplicity and clarity. Persuasion was initially predicted to play a minor role within the function of the SRF, and participant data showed nuanced support for this intervention function. Positive feelings regarding the SRF encouraged action. Finally, while Modelling had been considered a minor but important function of the SRF prior to the trial, no findings supported this.

#### c) Understand how the SRF worked through considering its behaviour change techniques

Finally, in relation to the SRF working as anticipated at the level of its BCTs (Table 3) we found broad support for many but not all the BCTs we had identified within the SRF. Our analysis of interview data suggested that the BCTs that played a particularly important role were from three main BCT groups: ‘Goals/planning’, ‘Monitoring & feedback’, and ‘Associations’. However, we found no empirical support for BCTs from ‘Shaping knowledge’ and ‘Antecedents’.

Within the ‘Goals/planning’ group, the specific BCT of ‘1.2 Problem solving’ was identified as an integral mechanism. “I think on occasion it’s helping us to go back to wards that patients have been on, where there’s overlaps that we’ve not recognised and intervene on those wards’, (Site 2, 694). Relevant ‘Monitoring & feedback’ BCTs were: ‘2.2 Feedback on behaviour’, ‘2.6 Biofeedback’, and ‘2.7 Feedback on outcomes of behaviour’. In relation to the ‘Associations’ group, there was strong support for ‘7.1 Prompts/cues’ being central to how the SRF worked in practice. Minimal support was found for ‘9.1 Credible source’. Analysis showed no support for ‘4.1 Instruction on how to perform a behaviour’ or ‘12.5 Adding objects to the environment’. Analysis also identified behaviour change elements which were not captured within the BCT taxonomy; particularly in relation to ‘Simplicity of design’.

### RQ3: How did the SRF work in practice?

We end with a narrative account reflecting our inductive thematic analysis. This provides a more contextualised account within which to place the previous analyses (RQ1 and RQ2). Despite the challenging backdrop of the COVID-19 pandemic, most participants shared the view that the SRF was acceptable, beneficial, and useful. Positive accounts of the SRF spanned three themes: its ease of understanding; its perceived efficacy and impact on IPC behaviours; and issues of assimilation into existing work.

#### Ease of understanding

Participants mostly supported the SRF enabling the easy understanding of insights from WGS. Participants described the SRF’s succinct and straightforward content as facilitating rapid action “at a glance” (Site 2). This sense of simplicity was often framed by implicit comparisons with other ways of communicating genetic information “it’s way more useful having a report like this than providing the phylogeny” (Site 1). Participants frequently cited the novel visualisation of transmission, along with the simple narrative conclusion, as the form’s greatest strengths. Participants strongly supported the idea that a key – and novel – component of the SRF was its use of visuals and plain English to communicate WGS insights:

> The visual timeline at the bottom of the report I think’s been particularly useful, especially useful when communicating the results to other staff members who are not, you know, so involved with the sequencing side of it … it’s really helpful to be able to show, you know, ward nurses: ‘look you’ve had this case, now it looks closely matched to this case, that was there five days ago, a week ago’ or whatever, and so yeah, I think that’s, that’s been really good. (Site 2)

In contrast, a very small minority of participants reported the SRF was difficult to use and interpret. Despite repeated assertions across the data from a variety of staff that the form’s simplicity rendered specialist training in interpreting its content unnecessary, a few staff did report residual confusion:

> I could not interpret [the SRF] at all. I purely looked for names and hospital numbers, but the actual information on it, and again that brings me back to having a little bit of background knowledge in relation to typing [I.e., genome sequencing] and how it works. I very much had to lean on my virology colleagues and micro [microbiology] colleagues who have more of a knowledge of typing. […] I definitely looked at it and went ‘I have no idea what that means, can somebody interpret that for me?’, which is a shame because I feel like it, I would love to look at it and ‘go, ah, that means that.’ […] But the actual physical form that came through to tell us, yeah I wouldn’t say I had a Scooby Doo [i.e., a clue], I’m afraid. […] We’d like more knowledge on that though. I felt a bit stupid I have to say. (Site 3)

#### Perceived efficacy and impact on IPC behaviours

In relation to perceived efficacy and impact, most participants shared the sense of the SRF’s particular value in objectively and rapidly tracking transmission pathways, and subsequently prompting IPC action:

> That was probably the most interesting part [looking at SRFs] because … you could actually see it working… so … when we were in a flow of “okay we’ve got this patient come through - this is the report”, and speaking to [HOCI PI], and then the nurses reacting to that, and making decisions based off of what we found in the reports, it was really interesting just to see the link between all of it, and just how it can help and it could help in the future. (Site 5)

The benefit of being presented with objective and actionable information was echoed by the majority of participants. The SRF’s provision of both timing and location of nosocomial cases was reported as crucial to staff’s capacity to “very quickly get a grip on what’s going on with this patient” (Site 3). This notion of the value of objective information – in the context of ongoing uncertainty – was echoed by many, evoking a sense of relief in knowing that “the sequence doesn’t lie” (Site 3). Gratitude for impartial clarity was oft repeated – participants almost unanimously agreed on the SRF providing “clear” and actionable information.

#### Issues of assimilation into existing work

In relation to assimilation into existing work, our analysis speaks to the complex context in which the SRF was used (i.e., the peak of the Alpha variant in Spring 2021). The SRF was assimilated in some sites and by some participants, but this was far from universal. On the one hand, where the SRF was seen to work in practice, it was able to be assimilated over time “there was a bit of a shift in the mindset of some of our infection control staff around ‘actually this could be very beneficial for us’” (Site 1). However, in other places and times, other factors constrained its assimilation. These related to the ‘flood’ of patients with COVID and the volume of patients with nosocomial infection, “it was simply too busy to do it, it would have been nice to have done it” (Site 5). There was a sense that there was a ‘goldilocks zone’ in which the SRF could work but that if there was too little or too much infection its ability to change IPC was limited:

> Although in the thick of it, I think we’re all thinking, actually, what, how realistic is it? I think, if you have one or two cases it’s more realistic. When you’re actually doing only what’s possible as opposed to what’s desirable, it maybe isn’t going to make a big difference. (Site 1)

The final issue that affected the assimilation of the SRF into existing work and practice related to a series of factors that extended beyond the SRF and related to what people did with the SRF and its results. These factors related to the dynamic organisational environments in which the SRF was introduced. Issues that affected the effective dissemination of the SRF’s insights included the ability to prioritise the SRF within the context of an unfolding hospital crisis, interpersonal processes such as inter- and intra-team dynamics, meetings and staff availability, software and innovation to enable team working on the SRFs within the COVID crisis. Elsewhere we focus exclusively on these implementation issues in more detail (Leiser et al., In preparation).

## Discussion

This paper presents a novel example of a behaviourally focused qualitative process evaluation. Using qualitative and behavioural analyses, we evaluated a simple form created to elicit changes in infection prevention and control behaviour to reduce nosocomial infections. Through a range of analytic approaches we provided a detailed qualitative sense of how the SRF worked in the context of the second wave of the UK COVID-19 pandemic. The novel and simple design offers a way for health psychologists to consider conducting future process evaluations using qualitative data and behavioural analyses but will not be suitable for all situations. Simultaneously, beyond what the paper achieves methodologically, our empirical findings enable us to consider what we have usefully learned about the SRF. In turn, this may help shape the future use of SRFs for other nosocomial infections. To our knowledge this paper is the first in the world to provide a focussed evaluation of SRFs within infectious disease.

In relation to our first research question (what were the putative active ingredients of the SRF?), our pre-trial analysis detailed a whole series of inter-related components and mechanisms. Core mechanisms were identified as ‘Knowledge’ and ‘Behavioural regulation’. In relation to the broad intervention functions of the SRF these were coded as ‘Education’ and ‘Enablement’ although ‘Persuasion’ and ‘Modelling’ were also considered relevant. Finally, several individual BCTs were thought to be deployed as a function of the SRF. These spanned a number of BCT groups – with the highest proportion of specific BCTs found within the Feedback & Monitoring group. For the first research question, the health psychology team were not involved within the work to design and optimise the SRF. Given the context (i.e., the COVID-19 pandemic), it was not possible to conduct systematic development work involving diverse HCPs. If time and resource had permitted, systematic and dedicated behaviourally informed qualitative work using focus groups and interviews and the use of approaches such as the think-out-loud approach (e.g., Van Someren et al., 1994) could all have enabled an optimal SRF to have gone to trial. This process would also have generated an agreed theorised account of the SRF’s putative content without the need for a post-hoc analysis of its content.

In relation to our second research question (was there evidence to support the SRF working as anticipated?), we found broad support for much of the content working as we imagined it would. Our analysis largely supported pre-trial conceptualisation of the SRF’s content and its function, with minimal exceptions. In relation to theoretical mechanisms, post-trial analysis strongly supported the putative importance of ‘Knowledge’ and ‘Behavioural regulation’, with both domains working as anticipated. Our analysis also showed that the relative importance of ‘Memory, attention, & decision-making processes’ and particular visual features were more important than we had anticipated before the trial. Analysis also provided support for three of the four intervention functions identified pre-trial: Education, Enablement, and Persuasion, however there was no support for Modelling. Participant data also showed strong support for previously identified BCTs. Our analysis found the strongest support for BCTs related mainly to feedback and monitoring: ‘Feedback on behaviour’, ‘Biofeedback’, and ‘Feedback on outcomes of behaviour’. In addition, ‘Problem solving’ and ‘Prompts/cues’ were both integral to the SRF and its function. However, disparity between pre-trial theorising and post-trial data was evident in the BCTs of ‘Instruction on how to perform a behaviour’ and ‘Adding objects to the environment’. Crucially, one key finding related to the importance of ‘Simplicity of design’ – something not captured within the BCTTv1 taxonomy but, nevertheless, evidently a fundamental component of the SRF. We explored the SRF content using the TDF, the BCW’s intervention functions and the BCTTv1. The process of exploring relative support for the SRF content from interview data using these three tools simultaneously was challenging at times. because much of our participant-led interview data seemed to speak simultaneously to the congruence of these three elements rather than solely to either TDF, intervention functions, or BCTs. Overall, we found identifying support for intervention functions more challenging than detecting palpable support for either the TDF domains or the BCTs. This does beg questions of whether using all three tools was actually needed. On reflection using the TDF and BCTTv1 alone would have elicited the same results. We might have had different results if we had used a more structured approach to data collection, however it may also be that distinguishing types of intervention content from the perspectives of those who use interventions is too challenging. In relation to our third research question (How did the SRF work in practice?), we found that the SRF was largely seen as useful at various levels; it was easy to understand, it appeared to work and have an impact on IPC behaviours, and in context-dependent ways some staff found it easy to assimilate into their existing work and professional practice. However, we now know from the wider quantitative trial results (Stirrup et al., 2022) that no statistically significant changes in weekly incidence of nosocomial SARS-CoV-2 were reported across the 14 trial sites, although in a sensitivity analysis, in 20.7% of nosocomial cases, when the SRF was returned within 5 days, there was an impact on IPC actions. These trial findings, in combination with the positive findings reported here, beg the question of why the SRF did not work as intended at changing the primary trial outcome. Our wider thematic analysis suggests that, beyond the form itself, the pathways to its implementation were particularly important. Elsewhere, we focus on these issues in greater detail, considering how best to support the implementation of the SRF to maximise use of its content (Leiser et al., in preparation).

### Strengths & Limitations

Strengths include the novel process evaluation design and use of health psychology approaches (i.e., TDF, BCW and BCTTv1) within an IPC intervention context. The comparison of pre- and post-trial analysis using these tools also provided added value. The fact we did find differences pre-and post-trial demonstrates the importance of theorising the intervention content before trial data collection. Gauging opinions on the SRF post-trial alone would have failed to provide as comprehensive an awareness of the underpinning mechanisms of the intervention, thereby lessening our capacity to evaluate the SRF’s content. Another strength is the study was its use of complementary deductive and inductive thematic analyses –generating an accessible narrative account of using the SRF but also to providing empirical evidence supporting the theorised mechanisms of the SRF. Lastly, collecting qualitative data from a wide range of different HCPs working with the SRF afforded an overall picture of experience that comprised both breadth and depth.

In relation to limitations, our reliance on interview-derived qualitative data alone raises questions about the veracity of our findings. We know that participant recall focuses on salience and feel confident that the findings reported here reflect what was most important and memorable to our participants. Equally, our use of tools to understand intervention content is becoming outdated. On-going work is revising the content of the BCTTv1 and indeed the way in which mechanisms can be theorised (https://theoryandtechniquetool.humanbehaviourchange.org/). Other limitations of the study relate to the span of our data collection. Interviews were carried out in only five out of fourteen trial sites from the wider study, which – although offering a varied and substantial sample – this doesn’t necessarily capture perspectives across the trial as a whole. However, these findings were shared and discussed in summer 2021 with a far broader range of staff involved in using the SRF from across all trial sites. Another limitation was the temporal time frame of data collection, largely taking place within weeks of each site delivering SRFs rapidly and at the peak of the Alpha variant within the COVID-19 pandemic. This presented unique challenges in collecting data on the way the SRF worked when our findings themselves suggest it took time for the SRF to embed and be understood. Embedding the rapid delivery of the SRF over a longer period and exploring longer term issues of implementation may give a richer source of understanding the SRF. Finally, the tremendous burden of COVID-19 on HCPs across the workforce cannot be overstated and may have influenced attitudes towards both the SRF itself, and participation within this study.

## Conclusion

This paper suggests both general and granular support for the SRF as an intervention that can change IPC behaviour. Empirical evidence, in the form of rich qualitative data, showed support for the previously theorised mechanisms of the SRF as an intervention to direct IPC behaviour to reduce nosocomial infection of SARS-CoV-2. The consolidation of both pre- and post-trial analysis provided a robust overview of how the SRF worked in practice, and also highlights its acceptability among the people who used it. However, to capitalise on SRF capacity to reduce nosocomial infection, future complementary work on embedding it into routine practice is required.

## Data Availability

Given the qualitative and sensitive nature of the data underpinning the analyses data are not available

## Acknowledgements

We would like to particularly acknowledge the support of NHS Greater Glasgow and Clyde Clinical Research Facility.

We also acknowledge the support of the independent members of the Joint Trial Steering Committee and Data Monitoring Committee (TSC-DMC): Prof Marion Koopmans (Erasmus MC), Prof Walter Zingg (University of Geneva), Prof Colm Bergin (Trinity College Dublin), Prof Karla Hemming (University of Birmingham), Prof Katherine Fielding (LSHTM). As well as TSC-DMC non-independent members: Prof Nick Lemoine (NIHR CRN), Prof Sharon Peacock (COG-UK). We would also thank members of COG-UK who have directly supported the study: Dr Ewan Harrison (Cambridge University), Dr Katerina Galai (PHE), Dr Francesc Coll (LSHTM), Dr Michael Chapman (HDR-UK), Prof Thomas Connor and team (Cardiff University), Prof Nick Loman and team (University of Birmingham). We also thank the COG-UK Consortium, and the UK National Institute for Health Research Clinical Research Network (NIHR CRN).

## Notes

**Conflict of interest statement** Paul Flowers – No competing interests declared Ruth Leiser – No competing interests declared Julie McLeod- No competing interests declared Fiona Mapp- No competing interests declared Oliver Stirrup - No competing interests declared Christopher JR Illingworth- No competing interests declared James Blackstone- No competing interests declared Judith Breuer- No competing interests declared

**Funding statement** This work was supported by funding from the Medical Research Council (MRC) part of UK Research & Innovation (UKRI), the National Institute of Health Research (NIHR) [grant code: MC_PC_19027], and Genome Research Limited, operating as the Wellcome Sanger Institute.

### Competing Interest Statement

The authors have declared no competing interest.

### Clinical Trial

NCT04405934

### Clinical Protocols

https://osf.io/preprints/socarxiv/ysm35/

### Funding Statement

This work was supported by funding from the Medical Research Council (MRC) part of UK Research & Innovation (UKRI), the National Institute of Health Research (NIHR) [grant code: MC_PC_19027], and Genome Research Limited, operating as the Wellcome Sanger Institute.

### Author Declarations

Ethical approval was given by Cambridge South Research Ethics Committee (20/EE/0118).

